# Evaluation of evidence for pathogenicity demonstrates that *BLK, KLF11* and *PAX4* should not be included in diagnostic testing for MODY

**DOI:** 10.1101/2021.09.17.21263728

**Authors:** Thomas W Laver, Matthew N Wakeling, Olivia Knox, Kevin Colclough, Caroline F Wright, Sian Ellard, Andrew T Hattersley, Michael N Weedon, Kashyap A Patel

## Abstract

Maturity Onset Diabetes of the Young (MODY) is an autosomal dominant form of monogenic diabetes, reported to be caused by variants in 16 genes. Concern has been raised about whether variants in *BLK* (MODY11), *KLF11* (MODY7) and *PAX4* (MODY9) cause MODY. We examined variant-level genetic evidence (co-segregation with diabetes and frequency in population) for published putative pathogenic variants in these genes and used burden testing to test gene-level evidence in a MODY cohort (n=1227) compared to population control (UK Biobank, n=185,898). For comparison we analysed well-established causes of MODY, *HNF1A* and *HNF4A*. The published variants in *BLK, KLF11* and *PAX4* showed poor co-segregation with diabetes (combined LOD scores ≤1.2), compared to *HNF1A* and *HNF4A* (LOD scores >9), and are all too common to cause MODY (minor allele frequency >4.95×10^−5^). Ultra-rare missense and protein-truncating variants (PTVs) were not enriched in a MODY cohort compared to the UK Biobank (PTVs *P*>0.05, missense *P*>0.1 for all three genes) while *HNF1A* and *HNF4A* were enriched (*P*<10^−6^). Sensitivity analyses using different population cohorts supported our results. Variant and gene-level genetic evidence does not support *BLK, KLF11* or *PAX4* as causes of MODY. They should not be included in MODY diagnostic genetic testing.

---

Maturity Onset Diabetes of the Young (MODY) is the most common subtype of monogenic diabetes. It is reported to be caused by heterozygous variants in 16 genes [1]. MODY accounts for approximately 3% of all diabetes cases under 30 years of age [2, 3]. The prevalence of MODY is estimated to be 108 cases per million [4]. An accurate genetic diagnosis is important for patients with MODY as it can determine the correct treatment [1, 5] and provides an accurate assessment of the risk of diabetes for future offspring. The advent of next generation sequencing has enabled a paradigm shift in genetic testing from focusing on single gene testing to gene panel tests for diseases [6]. While this can boost diagnostic yield, it does increase the risk of reporting variants in genes that are not a cause of MODY, as next generation sequencing enables testing of all genes regardless of evidence. An incorrect genetic diagnosis could result in stopping insulin in a patient with type 1 diabetes. It could lead to inappropriate testing of family members, causing increased anxiety in unaffected relatives and inflicting the psychological burden of having a genetic disease. Therefore, it is crucial that the gene panel only includes genes with robust aetiological evidence to prevent misdiagnosis of MODY.

*BLK, KLF11* and *PAX4* are listed on OMIM as MODY11, MODY7 and MODY9 but there is a need to revaluate whether variants in these genes do cause MODY. Variants in *BLK* and *PAX4* have been reported to cause MODY via haploinsufficiency [7, 8] while variants in *KLF11* were reported to cause the disease, potentially via a gain of function mechanism [9]. These studies were conducted more than 10 years ago, before the availability of variant frequency in large population cohorts [7-9]. *KLF11* and *PAX4* were identified based primarily on biological candidacy rather than the hypothesis-free genetic approach which is now considered to be the most robust method for gene discovery studies. The only *BLK* coding variant (p.A71T) reported to cause MODY was later found to be very common in the population raising doubt over the aetiological role of *BLK* [10]. Rarity of a variant in a large control population as well as enrichment of variants in that gene in a disease cohort compared to a control population have become crucial evidence to support the gene-disease association alongside familial co-segregation [11, 12].

Therefore, the aim of our study was to evaluate genetic evidence for variants in *BLK, KLF11* and *PAX4* as a cause of MODY. We evaluated the existing evidence for these genes and assessed the gene-disease association using a large MODY cohort and population cohorts. We demonstrate there is a lack of robust genetic evidence to support the aetiological role of variants in *BLK, KLF11* and *PAX4* for MODY.

## Research Design and Methods

### Study populations

#### MODY cohort

We included 1227 unrelated probands from the UK who were referred for genetic testing for MODY from routine clinical care to the Exeter Genomics Laboratory at the Royal Devon and Exeter Hospital. Cohort characteristics are provided in Supplementary Table 1. Informed consent was obtained from the probands or their parents/guardians and the study was approved by the North Wales ethics committee (17/WA/03).

#### UK Biobank

UK Biobank is a population-based cohort from the UK with deep phenotyping data and genetic data for around 500,000 individuals aged 40-70 years at recruitment [13, 14]. A subset of ∼200,000 DNA samples from UK Biobank participants underwent exome sequencing; this dataset was recently made available for research [15]. The UK Biobank resource was approved by the UK Biobank Research Ethics Committee and all participants provided written informed consent to participate.

#### GnomAD

We used GnomAD v.2.1.1 (141,456 individuals) and v3 (76,156 individuals) as alternative population controls in supplementary analyses. The detailed description of the cohort is previously published [16]. GnomAD v2.1.1 contains individuals with exome (n=125,748) and genome (n=15,708) data whereas v3 contains individuals with genome data.

### Genetic testing

#### MODY cohort

We undertook targeted next generation sequencing of *BLK, PAX4* and *KLF11* as well as *HNF1A* and *HNF4A* for probands suspected to have MODY, as previously described [6]. Targets were covered at a mean read depth of 460X per base and all bases had a mean coverage depth of at least 30 reads across the cohort. Variants were annotated against Genome Reference Consortium Human Build 37 (GRCh37) using Alamut Batch (Interactive Biosoftware, Rouen, France) using a Refseq transcript: *BLK* NM_001715.3, *KLF11* NM_003597.4, *PAX4* NM_001366110.1, *HNF1A* NM_000545.6, *HNF4A* NM_175914.4.

#### UK Biobank

We included 185,898 unrelated individuals from all ethnicities with exome sequencing data [17]. Detailed sequencing methodology for UK Biobank samples is provided by Szustakowski *et al*.[15] and is available at https://biobank.ctsu.ox.ac.uk/showcase/label.cgi?id=170. Briefly, exomes were captured with the IDT xGen Exome Research Panel v1.0 which targeted 39Mbp of the human genome with coverage exceeding on average 20x on 95.6% of sites. We included variants that had individual and variant missingness <10%, Hardy Weinberg Equilibrium p-value >10^−15^, minimum read depth of 7 for SNVs and 10 for indels, and at least one sample per site passed the allele balance threshold > 15% for SNVs and 20% for indels. Variants were called against Genome Reference Consortium Human Build 38 (GRCh38). We lifted the variants over to Build 37 (GRCh37) [18] to ensure compatibility with the variants from our MODY cohort then annotated using Alamut Batch (Interactive Biosoftware, Rouen, France) using a Refseq transcript: *BLK* NM_001715.3, *KLF11* NM_003597.4, *PAX4* NM_001366110.1, *HNF1A* NM_000545.6, *HNF4A* NM_175914.4.

#### GnomAD

The gnomAD consortium performed joint variant calling of the samples using a standardized BWA-Picard-GATK pipeline [16]. GnomAD was QCed and analysed using the Hail open-source framework for scalable genetic analysis (https://gnomad.broadinstitute.org/about). Variants in v2.1.1 were called against Genome Reference Consortium Human Build 37 (GRCh37), v3 against Build 38 (GRCh38). We lifted over v3 to Build 37 (GRCh37) then annotated all gnomAD variants using Alamut Batch (Interactive Biosoftware, Rouen, France) using a Refseq transcript: *BLK* NM_001715.3, *KLF11* NM_003597.4, *PAX4* NM_001366110.1, *HNF1A* NM_000545.6, *HNF4A* NM_175914.4. As gnomAD is an agglomeration of different sequencing projects, some genomic regions have low coverage in some samples therefore to control for this we removed the variants from both the MODY cohort and gnomAD cohorts if they were in a region of low coverage (≤10x coverage in ≤80% of samples) in either cohort or flagged as low quality in gnomAD.

#### Co-segregation analysis of putative pathogenic variants

We used author provided LOD (logarithm of the odds) scores where available for the first published variants in *BLK, PAX4, KLF11, HNF1A* and *HNF4A* which suggested the causal role of those variants in MODY. This was only available for *BLK* p.A71T [7]. If the LOD score was not provided, we calculated it based on the Gene Clinical Validity Curation Standard Operating Procedure [19]. We summed the LOD scores for multiple pedigrees where possible based on this guidance to calculate a combined LOD score. Using a binomial test we compared the observed proportion of family members with diabetes and a putative variant to the expected proportion of 0.5 if the variant was not associated with diabetes.

### Statistical analysis

For each analysis variant frequency was defined in the MODY cohort plus the control cohort combined. We compared the frequency of ultra-rare (allele count=1) protein truncating variants (PTVs) (essential splice site, stop gain and frameshift variants; excluding those in the last exon) and missense variants in each gene in the MODY cohort to the UK Biobank population cohort. We also provided the evidence of an association in terms of Bayesian false-discovery probabilities (BFDP) as previously described [20]. We replicated our analysis using two alternative population controls: gnomAD v2.1.1 (141,456 individuals) and gnomAD v3 (76,156 individuals) [16].

We used synonymous variants as a control to assess the difference in sequencing technologies and analysis pipeline. We also compared the frequency of rare variants (MAF<0.0001) and the frequency of all PTVs (no frequency filter) to test if there was an undue influence of ultra-rare variants due to differences in capture platforms. The most common *HNF1A* pathogenic variant is a frameshift variant (p.G292Rfs*25) in exon 4 due to a duplication of a C nucleotide. This variant is difficult to detect robustly in exome/genome sequencing data due to its location in a repetitive poly-C tract and the presence of a common variant that adds an additional 5’ C nucleotide to the tract (rs56348580 G>C, MAF=0.26). Since we were unable to perform confirmatory Sanger sequencing in the UK Biobank or GnomAD cohorts we excluded this variant from our analysis from all study cohorts.

We used Fisher’s exact test to assess variant enrichment in our MODY cohort and compute odds ratios with 95% confidence intervals. We used a threshold *P* value of 0.01 (0.05/5) as we tested 5 genes. We used Stata 16 (StataCorp, Texas, USA) for this analysis. BFDP was computed using a ‘gap’ R package. We used a prior probability of association of 0.99 for *HNF1A* and *HNF4A* to reflect the strong prior evidence for these genes and used 0.2 for *BLK, KLF11* and *PAX4* due to their probable disease association. We calculated the variance of the prior log(OR) as described by Wakefield[20], by assuming a 95% probability that the OR was less than 20 for *HNF1A* and *HNF4A*, and 3 for the other genes[20, 21]. We also explored different plausible priors as a sensitivity analysis.

## Results

### *BLK, KLF11* and *PAX4* variants had poor co-segregation in the published pedigrees

Variants that are highly penetrant causes of MODY would be expected to show strong co-segregation with the disease. To evaluate the genetic evidence of co-segregation with disease, we reviewed published pedigrees for putative variants in *BLK, KLF11* and *PAX4* causing MODY (Supplementary Table 2). We identified 1 *BLK*, 3 *KLF11* and 1 *PAX4* pedigrees with more than 3 individuals with variants to calculate LOD scores [7-9]. *KLF11* and *PAX4* variants showed poor co-segregation with diabetes in the families, with LOD scores of 1.2 and 0.6 respectively (Table 1). In line with low LOD scores, these variants were not associated with diabetes in family members in these pedigrees (P>0.5, Table 1). The *BLK* variant p.A71T also had a low LOD score of 1.16 and was modestly associated with diabetes in family members (*P*=0.02). In contrast, the variants reported in the first papers for *HNF1A* [22] and *HNF4A* [23], which are well-established causes of MODY, showed strong co-segregation with diabetes with combined LOD scores for the first reported variants of 9.63 and 15.05 respectively (Table 1).

**Table 1:**
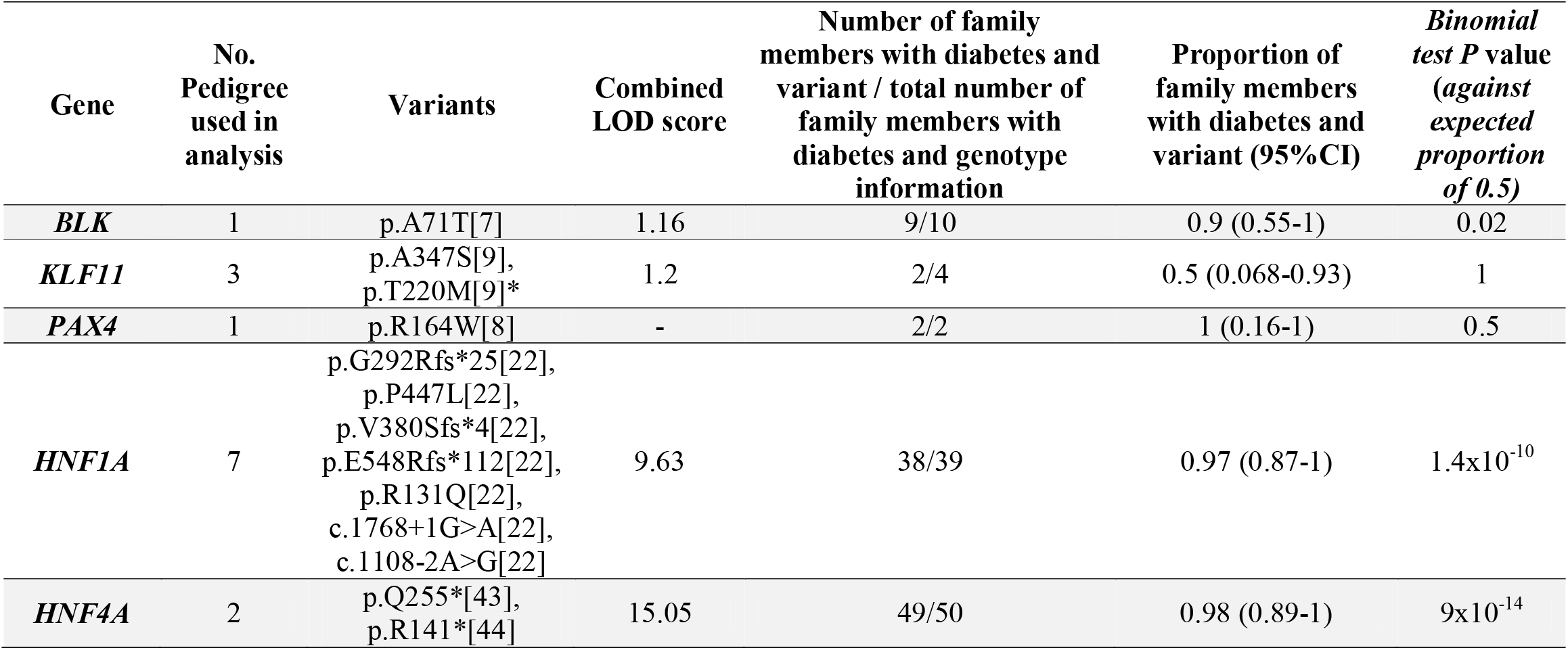
Co-segregation of *BLK, KLF11, PAX4* published variants with diabetes. Table shows the LOD scores and association of variants with diabetes in family members for variants where there were families with 3 or more people with the variant. We used author provided LOD scores where available for the first published variants which suggested the causal role of those variants in MODY. If the LOD score was not provided, we calculated it based on the Gene Clinical Validity Curation Standard Operating Procedure [19]. We summed the LOD score for each pedigree to calculate the combined LOD score. *two pedigrees with p.T220M were included in the combined LOD score calculation.

### Putative pathogenic variants in *BLK, KLF11* and *PAX4* are common in the population

The frequency of a putative pathogenic variant should not exceed the expected prevalence of the commonest variant in the commonest genetic subtype of the disease. MODY is estimated to have a population frequency of 1.08 per 10,000 [4]. We used the framework developed by Whiffin *et al* [24, 25] to calculate the maximum tolerated allele count in the population (gnomAD v2.1.1, n=141,456) for a putative pathogenic variant causing MODY. We used *HNF1A*, the most common cause of MODY, as a model to calculate the maximum tolerated allele count in the population. *HNF1A* accounts for 52% of MODY cases [4] and the most common mutation (p.G292Rfs*25) accounts for 19% of *HNF1A* cases [26]. At 50% penetrance the framework suggests that a pathogenic variant causing MODY should be present ≤3 times (frequency <2.1×10^−5^) in gnomAD v2.1.1 for *HNF1A*. As other genes will account for far fewer MODY cases, the putative pathogenic variants in *BLK, KLF11* and *PAX4* should be even rarer.

We looked at the frequency of variants in *BLK, KLF11* and *PAX4* that were reported to cause MODY before large scale population data was made available publicly in 2016 [27] (Table 2). The variants published since 2016 should have included the frequency of the variant in these databases as part of their screening process and thus would be expected to have only published rare variants (See Supplementary Table 2 for full list of HGMD variants in these genes).

**Table 2:** Population frequency of variants in *BLK, KLF11* and *PAX4* published as MODY causing. Allele frequency taken from gnomAD v2.1.1. The table provides coding variants reported before 2016 which are reported to cause MODY as the release of ExAC [27] that year meant variants published since then have had access to a large population control as part of their screening process. The *HNF1A* and *HNF4A* variants included here for comparison are those from the original papers used in the LOD score calculations in Table 1.

| Gene | Variant | Allele count /total alleles in gnomAD v2.1.1 | Allele frequency in gnomAD v2.1.1 | Allele count in ancestry with maximum frequency/total alleles in the ancestry | Maximum Allele frequency in a single ancestry in gnomadv2.1.1 (ancestry) | Reference for variants |
| --- | --- | --- | --- | --- | --- | --- |
| *BLK* | p.A71T | 3281/282812 | 0.012 | 420/10368 | 0.041 (Ashkenazi Jewish) | [7] |
| *KLF11* | p.Q62R | 25823/282778 | 0.091 | 1497/10370 | 0.144 (Ashkenazi Jewish) | [9] |
| | p.T220M | 1207/282762 | $4.27 \times 10^{-3}$ | 1098/24958 | 0.044 (African/African American) | [9] |
| | p.A347S | 36/282304 | $1.28 \times 10^{-4}$ | 17/35410 | $4.80 \times 10^{-4}$ (Latino/Admixed American) | [9] |
| *PAX4* | p.R31L | 105/250972 | $4.18 \times 10^{-4}$ | 102/30616 | 0.003 (South Asian) | [45] |
| | p.R164W | 14/282800 | $4.95 \times 10^{-5}$ | 3/24948 | $1.2 \times 10^{-4}$ (African/African American) | [8] |
| | p.R192H | 2214/282856 | $7.83 \times 10^{-3}$ | 2182/19946 | 0.109 (East Asian) | [8] |
| *HNF1A* | p.P447L | 3/249186 | $1.20 \times 10^{-5}$ | 1/20812 | $4.81 \times 10^{-5}$ (European Finnish) | [22] |
|  | p.V380Sfs*4 | 0 | 0 | 0 | 0 | [22] |
|  | p.E548Rfs*112 | 0 | 0 | 0 | 0 | [22] |
| | p.R131Q | 1/251390 | $3.98 \times 10^{-6}$ | 1/113698 | $8.80 \times 10^{-6}$ (European non-Finnish) | [22] |
|  | c.1768+1G>A | 0 | 0 | 0 | 0 | [22] |
|  | c.1108-2A>G | 0 | 0 | 0 | 0 | [22] |
| *HNF4A* | p.Q255* | 0 | 0 | 0 | 0 | [43] |
|  | p.R141* | 0 | 0 | 0 | 0 | [44] |

All putative MODY-causing variants in *BLK, KLF11* and *PAX4* published prior to 2016 were too common in the population to cause MODY. The allele count in gnomAD v2.1.1 was 4-8608 times higher than the maximum tolerable allele count for the commonest cause of MODY (Table 2). The least common was *PAX4* p.R164W which is seen 14 times in the whole of gnomAD v2.1.1 at a frequency of 4.95×10^−5^ but seen at higher frequency of 1.2×10^−04^ (3/24948) in the African/African American population. In contrast, the first reported variants in *HNF1A* and *HNF4A*, which were reported in the 1990s, are rare in the population with the most common (p.P447L) present 3 times in gnomAD v2.1.1 (1.20×10^−5^) (Table 2).

### Rare variants in *BLK, KLF11* and *PAX4* are not enriched in a MODY cohort

Having conducted variant level analyses on published variants in these genes we then carried out a gene level analysis to establish if other rare variants in these genes are likely to be pathogenic for MODY. To assess this, we carried out a gene burden test comparing the frequency of ultra-rare coding variants in a cohort of 1227 patients referred for MODY genetic testing with the frequency in the unrelated 185,898 exome-sequenced individuals from the population cohort UK Biobank (Table 3, Figure 1).

**Table 3:** Results of gene burden tests comparing the frequency of variants in *BLK, KLF11* and *PAX4* in a disease cohort to a population cohort. The frequency of ultra-rare (allele count=1) PTV and missense variants in a MODY cohort (n=1227) were compared to the frequency in the population cohort UK Biobank (n=185,898).

| Variant type | Gene | Allele count in MODY cohort | Allele frequency in MODY cohort | Allele count in Population cohort (UK biobank) | Allele frequency in Population cohort (UK Biobank) | Odds ratio (95%CI) | *P* | Prior Probability | Bayesian false-discovery probability (BFDP) |
| --- | --- | --- | --- | --- | --- | --- | --- | --- | --- |
| Ultra rare PTVs | *BLK* | 1 | 4.10x10<sup>-4</sup> | 14 | 3.77 x10<sup>-5</sup> | 10 (0.26-71) | 0.09 | 0.2 | 0.71 |
|  | *KLF11* | 0 | 0 | 15 | 4.03 x10<sup>-5</sup> | 0 (0-39) | 1 | 0.2 | 0.8 |
|  | *PAX4* | 0 | 0 | 6 | 1.61 x10<sup>-5</sup> | 0 (0-97) | 1 | 0.2 | 0.8 |
|  | *HNF1A* | 15 | 6.11x10<sup>-3</sup> | 3 | 8.07 x10<sup>-6</sup> | 762 (215-4108) | 1.37x10<sup>-30</sup> | 0.99 | 3.03 x 10<sup>-12</sup> |
|  | *HNF4A* | 3 | 1.22x10<sup>-3</sup> | 2 | 5.38 x10<sup>-6</sup> | 228 (26-2724) | 2.79x10<sup>-6</sup> | 0.99 | 6.74 x 10<sup>-5</sup> |
| Ultra rare Missense | *BLK* | 1 | 4.10x10<sup>-4</sup> | 141 | 3.79x10<sup>-4</sup> | 1 (0.03-6) | 0.6 | 0.2 | 0.82 |
|  | *KLF11* | 2 | 8.10x10<sup>-4</sup> | 122 | 3.28x10<sup>-4</sup> | 2.5 (0.3-9) | 0.2 | 0.2 | 0.78 |
|  | *PAX4* | 2 | 8.10x10<sup>-4</sup> | 84 | 2.26x10<sup>-4</sup> | 3.6 (0.43-13) | 0.1 | 0.2 | 0.70 |
|  | *HNF1A* | 21 | 8.56x10<sup>-3</sup> | 102 | 2.74x10<sup>-4</sup> | 31 (19-51) | 1.70x10<sup>-23</sup> | 0.99 | 1.46 x 10<sup>-40</sup> |
|  | *HNF4A* | 14 | 5.70x10<sup>-3</sup> | 83 | 2.23x10<sup>-4</sup> | 26 (13-46) | 4.41x10<sup>-15</sup> | 0.99 | 3.03 x 10<sup>-28</sup> |

**Figure 1:**
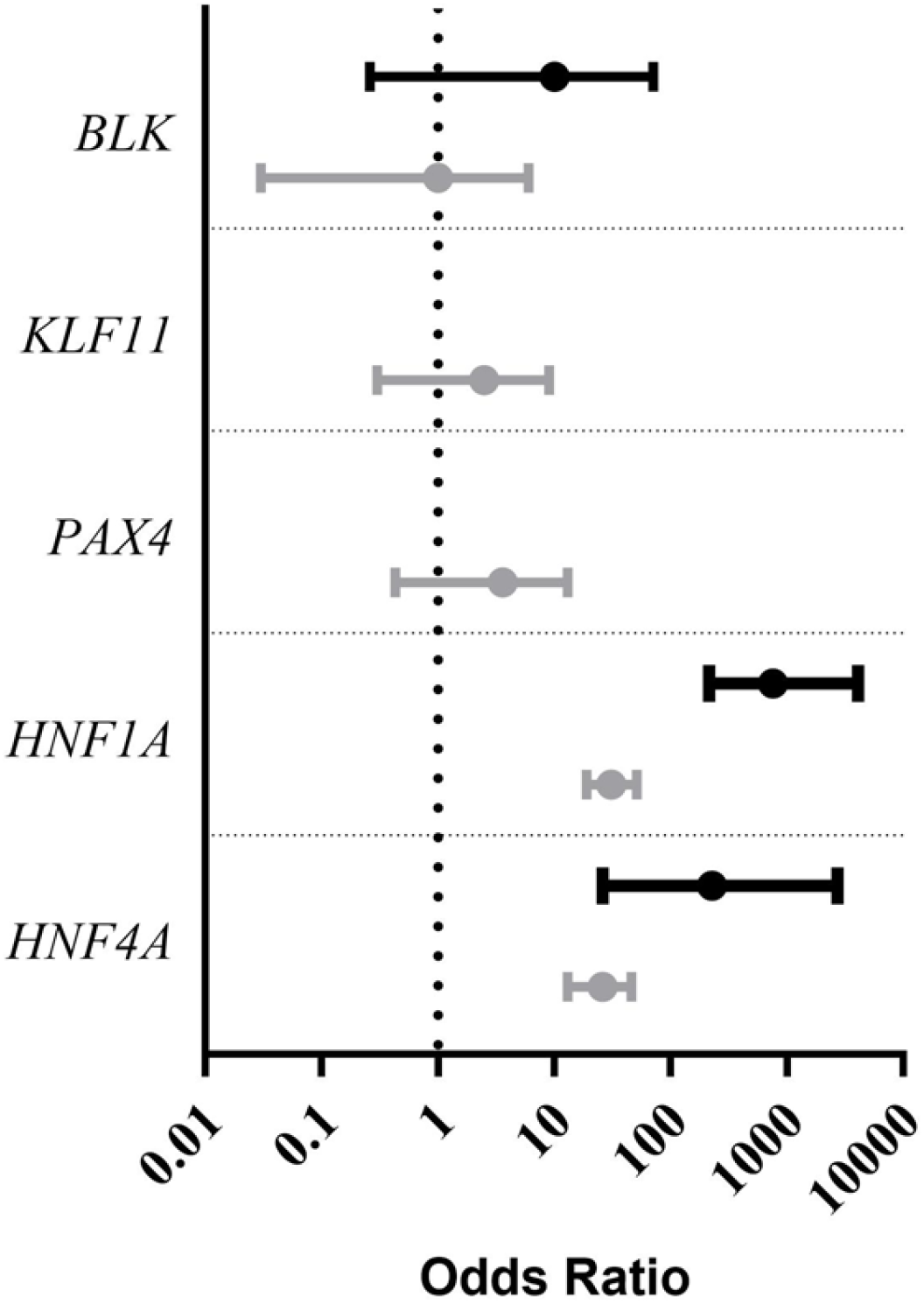
Graph showing the odds ratios for ultra-rare (allele count=1) protein truncating variants (PTVs) (black lines) and missense variants (grey lines) for MODY. Dotted line shows an odds ratio of 1. Bar not shown for *KLF11* and *PAX4* PTVs as odds ratio is 0 which cannot be plotted on a log axis.

Ultra-rare (allele count=1) PTV and missense variants in *BLK, KLF11* and *PAX4* are not enriched in our MODY cohort compared to the UK Biobank (all *P* values ≥0.09, Table 3). The bayesian false-discovery probability (BFDP) for ultra-rare PTV and missense variants was ≥0.70 for *BLK, KLF11* and *PAX4* (Table 3). The results of BFDP remained ≥0.37 using other plausible priors (Supplementary Table 3). In contrast variants in *HNF1A* and *HNF4A*, which are well established causative genes for MODY, were greatly enriched in our MODY cohort (all *P* values ≤2.79×10^−6^) with a very low BFDP (all ≤6.74 × 10^−5^).

### The lack of enrichment of rare variants in *BLK, KLF11* and *PAX4* is not due to technical artefacts

To ensure that our results are not due to differences in sequencing technologies or analysis pipelines between cases and controls, we performed a series of sensitivity analyses. Firstly, we analysed synonymous variant frequency in our MODY cohort and population control, and showed that the frequency of synonymous variants in all five genes was similar in our MODY cohort and the UK Biobank (all *P*>0.05, Supplementary Table 4).

Secondly, we replicated our gene burden analysis using gnomAD v.2.1.1 and v3 as two alternative population cohorts which were sequenced on different platforms (exome versus genome respectively) and with a different analysis pipeline versus the UK Biobank. Despite these differences, we found similar results with no enrichment in PTV or missense variants in *BLK, KLF11* or *PAX4* (Supplementary Tables 5 and 6).

Finally, to remove any undue influence of ultra-rare variants caused by differences in capture platforms, we performed a gene burden analysis for rare PTVs and missense variants (MAF <0.0001). We also compared the frequency of all PTVs in our MODY cohort and population control as all PTVs in these genes are considered to be pathogenic. These analyses showed similar results to our main analysis: rare PTVs and missense variants, and all PTVs, in *BLK, KLF11* and *PAX4* were not enriched in our MODY cohort whereas all these variant subsets in *HNF1A* and *HNF4A* showed great enrichment in our MODY cohort. (Supplementary Tables 7 and 8)

## Discussion

Variant and gene level genetic evidence presented in this study suggest that variants in *BLK, KLF11* and *PAX4* do not cause MODY. The lack of co-segregation of published MODY causing variants, presence in the population at high frequency and lack of enrichment of rare variants in a MODY cohort are consistent with these genes not causing MODY. The robustness of our approach is demonstrated by the results supporting the well-established causality of *HNF1A* and *HNF4A* variants.

Variants in *BLK, KLF11* and *PAX4* were reported to cause MODY more than 10 years ago, before large-scale variant population frequency became available [7-9]. Only small numbers of controls were available to rule out variants being present in the population.

*BLK* was first described in 2009 [7] by following up linkage to the 8p23 region [28] in 6 MODY families and identifying variants in *BLK* in three of the families. The frequency of the *BLK* variants was tested in 336 white control individuals and, for one variant, an additional 577 African American control individuals. *BLK* was identified via a linkage approach – it is possible that another candidate gene within the region of linkage is responsible for the disease in those families. Bonnefond *et al*. [10] found that the only non-synonymous variant in *BLK* reported to cause MODY was common in normoglycaemic individuals. This is the variant (p.A71T) that has a positive LOD score in the published pedigree; however as *BLK* was identified by linkage the LOD score would necessarily be positive regardless of the pathogenicity of the variant and, as also demonstrated by its frequency in gnomAD, the variant is clearly too common to cause MODY. No large MODY pedigrees with co-segregation have been described for *BLK* since the initial report. Non-coding variants in *BLK* were also reported to cause MODY [7], however, as our main cohorts consisted of targeted and exome sequencing data we were unable to investigate non-coding variants. It is unlikely that non-coding variants would be pathogenic given the lack of evidence for coding variants in *BLK* as a cause of MODY and that both coding and non-coding variants were proposed to cause the disease via loss of function.

*KLF11* was proposed as a cause of MODY via a candidate gene approach in 2005 [9]. The frequency of the reported *KLF11* variants was judged in only 313 normoglycemic individuals and 313 type 2 diabetes patients. Functional studies using Gal4 reporter assays suggested a possible mechanism of action for the variants via gain-of-function causing increased *KLF11* repression activity. If pathogenic variants in *KLF11* act via gain-of-function then we would not expect to see enrichment of PTVs in a MODY cohort, however we might expect to see enrichment of missense variants. We did not see enrichment of either type of variant and the previously identified *KLF11* variants are too common in the population to be disease causing. in patients from Thailand [8]. The variants were screened in a maximum of 344 non-diabetic controls. While their controls were from the same population as their cases, using data from gnomAD we now know that p.R192H is common in East Asians and both this variant and p.R164W are too common to cause MODY (p.R192H seen 2214 times in gnomAD v2.1.1 and p.R164W seen 14 times). Plengvidhya *et al*. [8] used luciferase reporter assays to show that p.R164W impairs the repressor activity of PAX4 on the insulin and glucagon promoters. However, they stated that the impairment was relatively small, thus it is possible that the reduction may be insufficient to result in a clinical phenotype. No large MODY pedigrees with co-segregation for a variant in *PAX4* have been described since the initial report.

Our study uses a large cohort of MODY cases and takes advantage of the availability of large population cohorts. The lack of enrichment for *BLK, KLF11* and *PAX4* PTV and missense variants in a MODY cohort compared to a population cohort is consistent with these genes not causing MODY. However, alternative explanations may be that the mechanism of action for these genes is not loss of function (as has been suggested for *KLF11* [9]) or they are an extremely rare cause of MODY. However, we did not see enrichment in missense variants (at either AC=1 or MAF<0.0001) suggesting that this is unlikely. In line with our results, gnomAD pLI and missense constraint scores for these genes are low, suggesting these genes are not under strong negative selection, in contrast to *HNF1A* and *HNF4A* which have high constraint scores. This data suggests that variants in these genes do not cause a rare monogenic disorder.

Variants in these genes could still be acting as polygenic risk factors for diabetes. Indeed, *PAX4* has been reported in the literature as a type 2 diabetes risk factor [29, 30]. However, PTVs and missense variants with MAF <1% in *BLK, KLF11* and *PAX4* are not associated with diabetes in the type 2 diabetes knowledge portal [31] and a recent large type 2 diabetes case control study did not find an association [32].

A limitation of our study is that by using publicly available population controls there were cross platform differences between cases and controls. This issue was mitigated by removing genomic positions with low coverage in one cohort from the other and by our sensitivity analyses: using synonymous variants as a negative control and testing alternative population control cohorts. Despite using a large MODY cohort there was still a relatively limited sample size of cases which could tend our gene burden tests of ultra-rare variants towards negative results. To ensure a lack of power was not determining the results we also used sensitivity analyses with MAF <0.001 and these did not suggest there was an association between *BLK, KLF11* or *PAX4* and MODY.

Our study results have important implications for genetic diagnostic laboratories worldwide who offer testing for MODY. Based on our results, we recommend that *BLK, KLF11* and *PAX4* should not be included in the gene panels for genetic testing for MODY and should not be reported as a cause of MODY. Studies are still reporting variants in these genes as a cause of MODY and they are routinely tested in clinical practice [33-37]. Our systematic review of the NCBI gene testing registry showed that 19 of 25 panels offered by diagnostic genetic laboratories still have at least one of these genes on their panel. Our study removes the ambiguity of the etiological role of these genes for MODY and provides the clearest results to date that refute their role as causative genes for MODY. Excluding these genes from diagnostic panels will prevent misdiagnosis of MODY and reduce workload for laboratories. The results from our study provide much needed evidence to gene curation efforts such as ClinGen and the Gene Curation Coalition to support the removal of these three genes from MODY genetic panels [38, 39]. We also strongly recommend that variants in *BLK, KLF11* and *PAX4* should be removed as a cause of MODY on databases such as HGMD [40], OMIM [41], ClinVar and panelapp [42] that are widely used by diagnostic laboratories and geneticists worldwide.

In conclusion, we present evidence from re-analysis of published variants in *BLK, KLF11* and *PAX4* that they are too common to cause MODY, have poor co-segregation with diabetes in those families and since the initial description no large MODY families with co-segregation of a variant have been published. We have then shown a lack of enrichment of rare variants in these genes in a MODY cohort compared to a population cohort providing evidence that rare variants in these genes do not cause MODY. Overall, the evidence does not support *BLK, KLF11* or *PAX4* as causes of MODY and they should not be included in diagnostic genetic testing.

## Data Availability

The GnomAD datasets are publicly available (https://gnomad.broadinstitute.org/). Data from the the UK Biobank is available to researchers by application to the access committee (https://www.ukbiobank.ac.uk/). For the MODY cohort the genotype data could be used to identify individuals and so cannot be made openly available. Access to data is open only through collaboration. Requests for collaboration will be considered following an application to the Genetic Beta Cell Research Bank (https://www.diabetesgenes.org/current-research/genetic-beta-cell-research-bank/). Contact by email should be directed to the Lead Nurse, Dr Bridget Knight.

## Acknowledgments

C.W, S.E., A.T.H., Mi.N.W. and K.A.P. designed the study. K.C., S.E., A.T.H. and K.A.P recruited patients and performed clinical phenotyping. T.W.L., Ma.N.W. and O.K. analysed the MODY cohort targeted sequencing data. Mi.N.W. and K.A.P. analysed the UK Biobank exome sequencing data. T.W.L, Ma.N.W., O.K., Mi.N.W. and K.A.P. carried out statistical analysis. T.W.L. and K.A.P. wrote the manuscript. All authors reviewed and approved the manuscript.

K.A.P. is the guarantor of this work and, as such, had full access to all the data in the study and takes responsibility for the integrity of the data and the accuracy of the data analysis. The authors have no conflicts of interest to declare.

This research has been conducted using the UK Biobank Resource under Application Number 49847. The current work was funded by Diabetes UK (19/0005994) and the MRC (grant no MR/T00200X/1). The authors would like to acknowledge use of the University of Exeter High-Performance Computing (HPC) facility in carrying out this work. T.W.L is supported by a lectureship funded by Research England’s Expanding Excellence in England (E3) fund. K.A.P has a Career Development fellowship funded by the Wellcome Trust (219606/Z/19/Z). A.T.H. was a recipient of a Wellcome Trust Senior Investigator award (grant WT098395/Z/12/Z) and is employed as a core member of staff within the NIHR funded Exeter Clinical Research Facility and is an NIHR senior investigator.

