## Supplemental tables for "Evaluation of evidence for pathogenicity demonstrates that *BLK, KLF11* and *PAX4* should not be included in diagnostic testing for MODY"

**Supplementary Tables**

**Supplementary Table 1: Characteristics of MODY cohort**

| Characteristics | MODY cohort |
| --- | --- |
| N | 1227 |
| Age of diagnosis of diabetes, years | 21 (14-30) |
| Female Sex | 58% |
| Age at recruitment, years | 30 (18-42) |
| BMI, kg/m^2^ | 25.1 (21.6-29.5) |
| Parents with diabetes | 72% |
| HbA1c, % | 7.5 (6.5-9.3) |
| HbA1c, mmol/mol | 58 (48-78) |
| On Insulin alone | 41% |
| On Insulin and other hypoglycaemic agents | 13% |
| On other hypoglycaemic agents alone | 47% |
| European ancestry (self-reported) | 84% |

**Supplementary Table 2: Population frequency of variants in BLK, KLF11 and PAX4 published as MODY causing**

Table showing coding variants where they were associated with MODY-like diabetes. Allele frequency taken from gnomAD v2.1.1. The *HNF1A* and *HNF4A* variants included here for comparison are those from the original papers used in the LOD score calculations in Table 1.

| Gene | Variant | Included in LOD score calculation | Allele count /total alleles in gnomAD v2.1.1 | Allele frequency in gnomAD v2.1.1 | Allele count in ancestry with maximum frequency/total alleles in the ancestry | Maximum Allele frequency in a single ancestry in gnomadv2.1.1 (ancestry) | Reference for variants |
| --- | --- | --- | --- | --- | --- | --- | --- |
| *BLK* | p.A71T | Yes | 3281/282812 | 0.012 | 420/10368 | 0.041 (Ashkenazi Jewish) | [1] |
| *KLF11* | p.Q62R | No | 25823/282778 | 0.091 | 1497/10370 | 0.144 (Ashkenazi Jewish) | [2] |
|  | p.T220M | Yes | 1207/282762 | 4.27x10^-03^ | 1098/24958 | 0.044 (African/African American) | [2] |
|  | p.A347S | Yes | 36/282304 | 1.28x10^-04^ | 17/35410 | 4.80x10^-04^  (Latino/Admixed American) | [2] |
| *PAX4* | p.R19Q | No | 4/251336 | 1.59x10^-05^ | 1/ 34592 | 2.89x10^-05^ (Latino/Admixed American) | [3] |
|  | p.R31L | No | 105/250972 | 4.18x10^-04^ | 102/30616 | 0.003 (South Asian) | [4] |
|  | p.R52C | No | 5/251274 | 1.99x10^-05^ | 1/18392 | 5.44x10^-05^ (East Asian) | [3] |
|  | p.A89V | No | 14/251416 | 5.57x10^-05^ | 11/113704 | 9.67x10^-05^ (European non-Finnish) | [3] |
|  | p.R97H | No | 8/251406 | 3.18x10^-05^ | 4/30616 | 1.31x10^-04^ (South Asian) | [5] |
|  | p.P142L | No | 5/251402 | 1.99x10^-05^ | 5/113720 | 4.4x10^-05^(European non-Finnish) | [3] |
|  | p.R164W | Yes | 14/282800 | 4.95x10^-05^ | 3/24948 | 1.2x10^-04^ (African/African American) | [6] |
|  | p.R192H | No | 2214/282856 | 7.83x10^-03^ | 2182/19946 | 0.109 (East Asian) | [6] |
|  | p.R192S | No | 783/282850 | 2.77x10^-03^ | 770/19950 | 0.039 (East Asian) | [7] |
|  | p.A198V | No | 0 | 0 | 0 | 0 | [8] |
| *HNF1A* | p.P447L | Yes | 3/249186 | 1.20x10^-05^ | 1/20812 | 4.81x10^-05^ (European Finnish) | [9] |
|  | p.V380Sfs*4 | Yes | 0 | 0 | 0 | 0 | [9] |
|  | p.E548Rfs*112 | Yes | 0 | 0 | 0 | 0 | [9] |
|  | p.R131Q | Yes | 1/251390 | 3.98x10^-06^ | 1/113698 | 8.80x10^-06^ (European non-Finnish) | [9] |
|  | c.1768+1G>A | Yes | 0 | 0 | 0 | 0 | [9] |
|  | c.1108-2A>G | Yes | 0 | 0 | 0 | 0 | [9] |
| *HNF4A* | p.Q255* | Yes | 0 | 0 | 0 | 0 | [10] |
|  | p.R141* | Yes | 0 | 0 | 0 | 0 | [11] |

**Supplementary Table 3: Bayesian false discovery probability at range of prior probability for enrichment of variant in MODY cohort versus UK Biobank**

| Variant type | Gene | BFDP at 0.5 | BFDP at 0.2 | BFDP at 0.1 |
| --- | --- | --- | --- | --- |
| Ultra-rare PTV | *BLK* | 0.38 | 0.71 | 0.85 |
|  | *KLF11* | 0.50 | 0.80 | 0.90 |
|  | *PAX4* | 0.50 | 0.80 | 0.90 |
|  | *HNF1A* | 3.00 x 10^-10^ | 1.20 x 10^-09^ | 2.70 x 10^-09^ |
|  | *HNF4A* | 0.01 | 0.03 | 0.06 |
| Ultra-rare missense | *BLK* | 0.54 | 0.82 | 0.91 |
|  | *KLF11* | 0.46 | 0.78 | 0.89 |
|  | *PAX4* | 0.37 | 0.70 | 0.84 |
|  | *HNF1A* | 1.45 x 10^-38^ | 5.80 x 10^-38^ | 1.31 x 10^-37^ |
|  | *HNF4A* | 3.00 x 10^-26^ | 1.20 x 10^-25^ | 2.70 x 10^-25^ |
| Rare PTV | *BLK* | 0.26 | 0.58 | 0.76 |
|  | *KLF11* | 0.15 | 0.41 | 0.61 |
|  | *PAX4* | 0.27 | 0.60 | 0.77 |
|  | *HNF1A* | 1.82 x 10^-30^ | 7.28 x 10^-30^ | 1.64 x 10^-29^ |
|  | *HNF4A* | 7.94 x 10^-05^ | 3.18 x 10^-04^ | 7.14 x 10^X04^ |
| Rare missense | *BLK* | 0.63 | 0.87 | 0.94 |
|  | *KLF11* | 0.66 | 0.88 | 0.95 |
|  | *PAX4* | 0.50 | 0.80 | 0.90 |
|  | *HNF1A* | 2.61 x 10^-34^ | 1.05 x 10^-33^ | 2.35 x 10^-33^ |
|  | *HNF4A* | 4.45 x 10^-22^ | 1.78 x 10^-21^ | 4.01 x 10^-21^ |
| All PTV | *BLK* | 0.26 | 0.58 | 0.76 |
|  | *KLF11* | 0.15 | 0.41 | 0.61 |
|  | *PAX4* | 0.41 | 0.74 | 0.86 |
|  | *HNF1A* | 2.10 x 10^-30^ | 8.41 x 10^-30^ | 1.89 x 10^-29^ |
|  | *HNF4A* | 7.94 x 10^-05^ | 3.18 x 10^-04^ | 7.14 x 10^-04^ |

**Supplementary Table 4: Gene burden test for synonymous variants in MODY cohort and UK Biobank**

The frequency of ultra-rare (allele count=1) synonymous variants in a MODY cohort n=1227 were compared to the frequency in the UK Biobank n=185,898.

*excluding two synonymous variants in *HNF1A* that were on a haplotype with a pathogenic PTV

| Variant type | Gene | Allele count in MODY cohort | Allele frequency in MODY cohort | Allele count in Population cohort (UK biobank) | Allele frequency in Population cohort (UK Biobank) | Odds ratio (95%CI) | *P* value |
| --- | --- | --- | --- | --- | --- | --- | --- |
| Ultra-rare synonymous | *BLK* | 0 | 0 | 61 | 1.64x10^-04^ | 0 (0-9.5) | 1 |
|  | *KLF11* | 0 | 0 | 55 | 1.48x10^-04^ | 0 (0-11) | 1 |
|  | *PAX4* | 0 | 0 | 38 | 1.02x10^-04^ | 0 (0-15) | 1 |
|  | *HNF1A* | 3 | 1.22x10^-03^ | 47 | 1.26x10^-04^ | 9.7 (1.9-30) | 0.004 |
|  | *HNF1A** | 1 | 4.07x10^-04^ | 47 | 1.26x10^-04^ | 3.2 (0.08-19) | 0.27 |
|  | *HNF4A* | 0 | 0 | 38 | 1.02x10^-04^ | 0 (0-15) | 1 |

**Supplementary Table 5: Gene burden test using gnomAD v2.1.1 as an alternative population control cohort**

The frequency of ultra-rare (allele count=1) PTV, missense and synonymous variants in a MODY cohort n=1227 were compared to the frequency in the GnomAD v2.1.1 n=141,456.

*excluding two synonymous variants in *HNF1A* that were on a haplotype with a pathogenic PTV

| Variant type | Gene | Allele count in MODY cohort | Allele frequency in MODY cohort | Allele count in Population cohort (GnomAD v2.1.1) | Allele frequency in Population cohort (GnomAD v2.1.1) | Odds ratio (95%CI) | *P* value | Prior Probability | Bayesian false discovery probability (BFDP) |
| --- | --- | --- | --- | --- | --- | --- | --- | --- | --- |
| Ultra-rare PTVs | *BLK* | 1 | 4.07x10^-04^ | 28 | 1.12x10^-04^ | 3.7 (0.089-22) | 0.2 | 0.2 | 0.78 |
|  | *KLF11* | 0 | 0 | 29 | 1.15x10^-04^ | 0 (0-14) | 1 | 0.2 | 0.80 |
|  | *PAX4* | 0 | 0 | 6 | 2.40x10^-05^ | 0 (0-65) | 1 | 0.2 | 0.80 |
|  | *HNF1A* | 13 | 5.30x10^-03^ | 3 | 1.21x10^-05^ | 441 (121-2415) | 3.93x10^-24^ | 0.99 | 3.36E-03 |
|  | *HNF4A* | 3 | 1.22x10^-03^ | 6 | 2.41x10^-05^ | 51 (8.2-238) | 1.00x10^-04^ | 0.99 | 0.07 |
| Ultra-rare Missense | *BLK* | 2 | 8.15x10^-04^ | 187 | 7.85x10^-04^ | 1 (0.12-3.8) | 0.7 | 0.2 | 0.84 |
|  | *KLF11* | 1 | 4.07x10^-04^ | 191 | 7.60x10^-04^ | 0.54 (0.013-3.0) | 1 | 0.2 | 0.82 |
|  | *PAX4* | 4 | 1.63x10^-03^ | 118 | 4.72x10^-04^ | 3.5 (0.93-9.1) | 0.03 | 0.2 | 0.48 |
|  | *HNF1A* | 18 | 7.33x10^-03^ | 151 | 6.07x10^-04^ | 12 (7-20) | 1.20x10^-13^ | 0.99 | 5.58E-16 |
|  | *HNF4A* | 10 | 4.07x10^-03^ | 105 | 4.21x10^-04^ | 9.7 (4.5-19) | 2.25x10^-07^ | 0.99 | 8.97E-07 |
| Ultra-rare Synonymous | *BLK* | 0 | 0 | 90 | 3.59x10^-04^ | 0(0-4.4) | 1 | NA | NA |
|  | *KLF11* | 1 | 4.07x10^-04^ | 83 | 3.30x10^-04^ | 1.2(0.031-7.1) | 0.6 | NA | NA |
|  | *PAX4* | 0 | 0 | 53 | 2.12x10^-04^ | 0(0-7.4) | 1 | NA | NA |
|  | *HNF1A* | 4 | 1.63x10^-03^ | 91 | 3.66x10^-04^ | 4.5(1.2-12) | 0.014 | NA | NA |
|  | *HNF1A** | 2 | 8.15x10^-04^ | 91 | 3.66x10^-04^ | 2.2(0.27-8.3) | 0.23 | NA | NA |
|  | *HNF4A* | 0 | 0 | 66 | 2.65x10^-04^ | 0(0-5.9) | 1 | NA | NA |

**Supplementary Table 6: Gene burden test using gnomAD v3 as an alternative population control cohort**

The frequency of ultra-rare (allele count=1) PTV and missense variants in a MODY cohort n=1227 were compared to the frequency in the GnomAD v3 n=76,156.

*excluding two synonymous variants in *HNF1A* that were on a haplotype with a pathogenic PTV

| Variant type | Gene | Allele count in MODY cohort | Allele frequency in MODY cohort | Allele count in Population cohort (GnomAD v3) | Allele frequency in Population cohort (GnomAD v3) | Odds ratio (95%CI) | *P* value | Prior Probability | Bayesian false discovery probability (BFDP) |
| --- | --- | --- | --- | --- | --- | --- | --- | --- | --- |
| Ultra-rare PTVs | *BLK* | 1 | 0.00041 | 22 | 0.000154 | 2.7 (0.064-16) | 0.3 | 0.2 | 0.80 |
|  | *KLF11* | 2 | 0.00081 | 8 | 5.58x10^-05^ | 15 (1.5-73) | 0.012 | 0.2 | 0.44 |
|  | *PAX4* | 1 | 0.00041 | 5 | 3.49x10^-05^ | 12 (0.25-104) | 0.1 | 0.2 | 0.73 |
|  | *HNF1A* | 17 | 0.00693 | 1 | 6.98x10^-06^ | 999 (156-41761) | 1.18x10^-29^ | 0.99 | 0.71 |
|  | *HNF4A* | 3 | 0.00122 | 3 | 2.09x10^-05^ | 58 (7.8-437) | 1.00x10^-04^ | 0.99 | 0.44 |
| Ultra-rare Missense | *BLK* | 2 | 8.15x10^-04^ | 154 | 1.07x10^-03^ | 0.76 (0.09-2.8) | 1 | 0.2 | 0.83 |
|  | *KLF11* | 3 | 1.22x10^-03^ | 125 | 8.72x10^-04^ | 1.4 (0.29-4.2) | 0.5 | 0.2 | 0.84 |
|  | *PAX4* | 2 | 8.15x10^-04^ | 70 | 4.89x10^-04^ | 1.7 (0.2-6.3) | 0.3 | 0.2 | 0.81 |
|  | *HNF1A* | 24 | 9.78x10^-03^ | 97 | 6.77x10^-04^ | 15 (8.9-23) | 6.87x10^-19^ | 0.99 | 9.16E-29 |
|  | *HNF4A* | 15 | 6.11x10^-03^ | 64 | 4.47x10^-04^ | 14 (7.3-24) | 4.67x10^-12^ | 0.99 | 6.91E-16 |
| Ultra-rare Synonymous | *BLK* | 1 | 4.07x10^-04^ | 44 | 3.07x10^-04^ | 1.3 (0.33-7.8) | 0.53 | NA | NA |
|  | *KLF11* | 1 | 4.07x10^-04^ | 68 | 4.75x10^-04^ | 0.86 (0.021-4.9) | 1 | NA | NA |
|  | *PAX4* | 0 | 0 | 30 | 2.09x10^-04^ | 0 (0-7.5) | 1 | NA | NA |
|  | *HNF1A* | 5 | 2.04x10^-03^ | 60 | 4.19x10^-04^ | 4.9 (1.5-12) | 0.0048 | NA | NA |
|  | *HNF1A** | 3 | 1.22x10^-03^ | 60 | 4.19x10^-04^ | 2.9 (0.59-9) | 0.09 | NA | NA |
|  | *HNF4A* | 0 | 0 | 52 | 3.63x10^-04^ | 0 (0-4.3) | 1 | NA | NA |

**Supplementary Table 7: Gene burden test for rare variants (MAF<0.0001) in MODY cohort and UK Biobank.** The frequency of rare (MAF<0.0001) PTV and missense variants in a MODY cohort n=1227 were compared to the frequency in the UK Biobank n=185,898.

*excluding two synonymous variants in *HNF1A* that were on a haplotype with a pathogenic PTV

| Variant type | Gene | Allele count in MODY cohort | Allele frequency in MODY cohort | Allele count in Population cohort (UK biobank) | Allele frequency in Population cohort (UK Biobank) | Odds ratio (95%CI) | *P* value | Prior Probability | Bayesian false discovery probability (BFDP) |
| --- | --- | --- | --- | --- | --- | --- | --- | --- | --- |
| Rare PTVs | *BLK* | 3 | 0.0012 | 124 | 0.0003335 | 3.7 (0.75-11) | 0.05 | 0.2 | 0.58 |
|  | *KLF11* | 2 | 0.0008 | 33 | 8.876x10^-05^ | 9.2 (1.1-36) | 0.02 | 0.2 | 0.41 |
|  | *PAX4* | 2 | 0.0008 | 55 | 0.0001479 | 5.5 (0.65-21) | 0.05 | 0.2 | 0.60 |
|  | *HNF1A* | 22 | 0.009 | 8 | 2.152x10^-05^ | 420 (180-1092) | 4.71x10^-42^ | 0.99 | 1.84 x 10^-32^ |
|  | *HNF4A* | 3 | 0.0012 | 4 | 1.076x10^-05^ | 114 (17-673) | 9.66x10^-06^ | 0.99 | 8.02 x 10^-07^ |
| Rare Missense | *BLK* | 7 | 0.0029 | 1245 | 0.0033 | 0.85 (0.34-1.8) | 0.9 | 0.2 | 0.87 |
|  | *KLF11* | 9 | 0.0037 | 1236 | 0.0033 | 1.1 (0.5-2.1) | 0.7 | 0.2 | 0.88 |
|  | *PAX4* | 8 | 0.0033 | 749 | 0.0020 | 1.6 (0.7-3.2) | 0.2 | 0.2 | 0.80 |
|  | *HNF1A* | 45 | 0.0183 | 1006 | 0.0027 | 6.9 (5-9.3) | 1.98x10^-22^ | 0.99 | 2.64 x 10^-36^ |
|  | *HNF4A* | 26 | 0.0106 | 583 | 0.0016 | 6.9 (4.4-10) | 1.42x10^-13^ | 0.99 | 4.50 x 10^-24^ |
| Rare Synonymous | *BLK* | 6 | 2.44x10^-03^ | 756 | 2.03x10^-03^ | 1.2 (0.44-2.6) | 0.65 | NA | NA |
|  | *KLF11* | 1 | 4.07x10^-04^ | 514 | 1.38x10^-03^ | 0.29 (0.0074-1.6) | 0.27 | NA | NA |
|  | *PAX4* | 0 | 0 | 181 | 4.87x10^-04^ | 0 (0-3.2) | 0.64 | NA | NA |
|  | *HNF1A* | 10 | 4.07x10^-03^ | 1014 | 2.73x10^-03^ | 1.5 (0.71-2.8) | 0.24 | NA | NA |
|  | *HNF1A** | 8 | 3.26x10^-03^ | 1014 | 2.73x10^-03^ | 1.2 (0.51-2.4) | 0.56 | NA | NA |
|  | *HNF4A* | 5 | 2.04x10^-03^ | 581 | 1.56x10^-03^ | 1.3 (0.42-3.1) | 0.44 | NA | NA |

**Supplementary Table 8: Gene burden test for all PTVs excluding last exon in MODY cohort and UK Biobank**

The frequency of PTV variants in a MODY cohort n=1227 were compared to the frequency in the UK Biobank n=185,898.

| Variant type | Gene | Allele count in MODY cohort | Allele frequency in MODY cohort | Allele count in Population cohort (UK biobank) | Allele frequency in Population cohort (UK Biobank) | Odds ratio (95%CI) | *P* value | Prior Probability | Bayesian false discovery probability (BFDP) |
| --- | --- | --- | --- | --- | --- | --- | --- | --- | --- |
| PTVs | *BLK* | 3 | 0.00122 | 124 | 0.000334 | 3.7 (0.75-11) | 0.052 | 0.2 | 0.58 |
|  | *KLF11* | 2 | 0.00081 | 33 | 8.88x10^-05^ | 9.2 (1.1-36) | 0.02 | 0.2 | 0.41 |
|  | *PAX4* | 2 | 0.00081 | 94 | 0.000253 | 3.2 (0.38-12) | 0.1 | 0.2 | 0.74 |
|  | *HNF1A* | 22 | 0.00896 | 8 | 2.15x10^-05^ | 420 (180-1093) | 4.71x10^-42^ | 0.99 | 2.12x10^-32^ |
|  | *HNF4A* | 3 | 0.00122 | 4 | 1.08x10^-05^ | 114 (17-673) | 9.66x10^-06^ | 0.99 | 8.02x10^-07^ |
